# Interactions between seasonal human coronaviruses and implications for the SARS-CoV-2 pandemic: A retrospective study in Stockholm, Sweden, 2009-2020

**DOI:** 10.1101/2020.10.01.20205096

**Authors:** Robert Dyrdak, Emma B. Hodcroft, Martina Wahlund, Richard A. Neher, Jan Albert

**Affiliations:** Department of Clinical Microbiology, Karolinska University Hospital, Stockholm, Sweden; Department of Microbiology, Tumor and Cell Biology, Karolinska Institutet, Stockholm, Sweden; Biozentrum, University of Basel, Basel, Switzerland; Swiss Institute of Bioinformatics, Basel, Switzerland; Department of Medicine, Infectious Diseases Unit, Center for Molecular Medicine, Karolinska Institutet, Stockholm, Sweden

## Abstract

**Objectives:** The four seasonal coronaviruses 229E, NL63, OC43, and HKU1 are frequent causes of respiratory infections and show annual and seasonal variation. Increased understanding about these patterns could be informative about the epidemiology of SARS-CoV-2.

**Methods:** Results from PCR diagnostics for the seasonal coronaviruses, and other respiratory viruses, were obtained for 55,190 clinical samples analysed at the Karolinska University Hospital, Stockholm, Sweden, between 14 September 2009 and 2 April 2020.

**Results:** Seasonal coronaviruses were detected in 2,130 samples (3.9%) and constituted 8.1% of all virus detections. OC43 was most commonly detected (28.4% of detections), followed by NL63 (24.0%), HKU1 (17.6%), and 229E (15.3%). The overall fraction of positive samples was similar between seasons, but at species level there were distinct biennial alternating peak seasons for the *Alphacoronaviruses*, 229E and NL63, and the *Betacoronaviruses*, OC43 and HKU1, respectively. The *Betacoronaviruses* peaked earlier in the winter season (Dec-Jan) than the *Alphacoronaviruses* (Feb-Mar). Coronaviruses were detected across all ages, but diagnostics were more frequently requested for paediatric patients than adults and the elderly. OC43 and 229E incidence was relatively constant across age strata, while that of NL63 and HKU1 decreased with age.

**Conclusions:** Both the *Alphacoronaviruses* and *Betacoronaviruses* showed alternating biennial winter incidence peaks, which suggests some type of immune mediated interaction. Symptomatic reinfections in adults and the elderly appear relatively common. Both findings may be of relevance for the epidemiology of SARS-CoV-2.

## Introduction

The world is seeing a pandemic by a new coronavirus, SARS-CoV-2, which is the cause of the disease COVID-19. The pandemic has unprecedented effects on societies globally. Although SARS-CoV-2 is new to mankind, six other coronaviruses are known to infect humans. This includes the four “seasonal” coronaviruses (CoVs): 229E, NL63, OC43 and HKU1, which are found globally and in immunocompetent hosts usually cause mild to moderate upper-respiratory tract disease [1-5]. In contrast, infections with two other coronaviruses, SARS-CoV and MERS-CoV, have severe clinical presentation and substantial mortality [6]. The human coronaviruses are found in two genera of the subfamily orthocoronavirinae. The genus *Alphacoronavirus* includes 229E and NL63, whereas the genus *Betacoronavirus* includes OC43 and HKU1 (subgenus Embecovirus), SARS-CoV and SARS-CoV-2 (subgenus Sarbecovirus), and MERS-CoV (subgenus Merbecovirus) [7].

Here we use results from routine clinical diagnostic tests on approximately 55,000 patient samples to analyze the epidemiology of the seasonal CoVs in Stockholm, Sweden. Better knowledge about the epidemiology of the seasonal CoVs could potentially inform about the epidemiology of SARS-CoV-2.

## Methods

### Samples and metadata

Data from routine diagnostics were obtained from the laboratory information system at the Department of Clinical Microbiology, Karolinska University Hospital, Stockholm, Sweden. The majority of samples analyzed were obtained from the Stockholm Region (2.2 million inhabitants), where the laboratory provides diagnostic services to six of seven major hospitals, and outpatient care to approximately to half of the population. PCR diagnostics for the seasonal CoVs were performed using accredited in-house assays [8] or the Allplex Respiratory Panels 2 and 3 (Seegene Inc., Seoul (South Korea)), see supplement for details on virus diagnostics. The associated metadata contained the laboratory results, date of sample collection, specimen type, age and sex of the patient. The study was reviewed and approved by the Swedish Ethical Review Authority (registration no. 2020-03001).

### Statistical analyses

Statistical analyses were performed using Stata version 15.1 (StataCorp LLC, College Station (TX, USA)). Point estimates and 95% confidence intervals (CIs) of proportions and odds ratios (ORs) were calculated using logistic regression with 200 bootstrap replications. To avoid selection bias, odds ratio for co-detections were calculated across the subset of samples with complete records and at least detection of one respiratory pathogen [9].

## Results

### Basic characteristics of samples and results from CoVs diagnostics

During 14 September 2009 until 2 April 2020, a total of 135,922 samples were obtained for respiratory virus diagnostics. Of these, 55,190 samples had been analyzed for an extended virus panel which included the four seasonal CoVs. There were 2153 detections of seasonal CoVs in 2,130 of the 55,190 samples (3.9%; 95% CI: 3.7%-4.0%), representing 8.1% (95% CI: 7.7%-8.4%) of all virus detections. The annual number of CoV-positive samples increased over the study period as a result of increased testing, but the fraction of positive samples decreased (Figure 1A and B, respectively). Among the 2,130 CoV-positive samples the most common species detected was OC43 (28.4%), followed by NL63 (24.0%), HKU1 (17.6%), and 229E (15.3%). In addition, 15.9% of the CoV-positive samples were positive for OC43/HKU1 in the Allplex assay, which does not distinguish between the two. In 23 samples (1.1% of all positive samples), two different species of CoVs were detected. Information about the sex of the patients was available for 55,102 samples (99.8%). CoV-positive results were significantly more common in men (55.1% of positive samples, 95% CI: 54.7%-55.5%, OR 1.16; 95% CI 1.06-1.26).

**FIG. 1.**
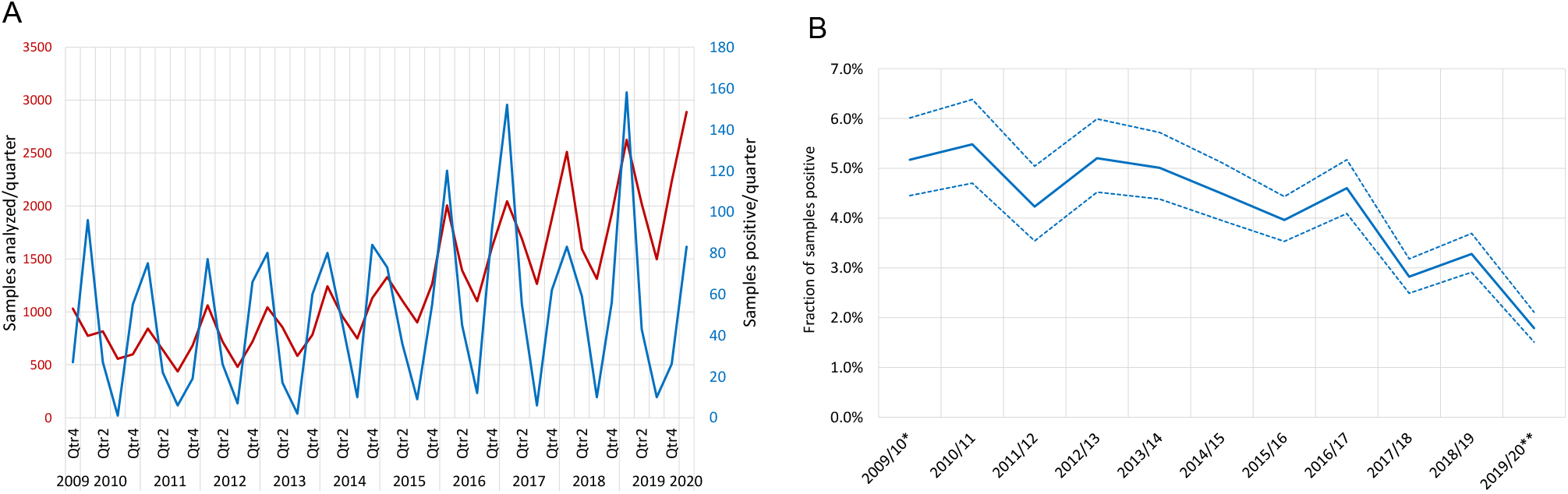
Samples positive for coronaviruses. (A) The number of samples analyzed per quarter from 1 October 2009 to 31 March 2020 (red line, *n* = 55,017), and samples being positive for any of the endemic coronaviruses (blue line, *n* = 2,128). (B) The fraction of samples positive per season for any of the seasonal coronaviruses. *, from 14 Sept until 31 Dec 2009; **, from 1 Jan until 2 April 2020.

### Alternating biennial incidence and peak incidence month of seasonal CoVs

The number of samples analyzed for CoVs (and other respiratory viruses) showed seasonal variation with peaks in the winters (Figure 1A). The number of CoV positive samples showed even more pronounced winter peaks. The overall fraction of samples positive for any species did not show any marked season-to-season variation across the duration of the study (Figure 1B). This is in stark contrast to the incidence of detection of the four CoV species, which all showed distinct biennial patterns (Figure 2). Interestingly, a pattern of alternating peak seasons was observed for the two *Alphacoronaviruses*, 229E and NL63. This pattern was even more striking for the two *Betacoronaviruses*, OC43 and HKU1. Due to the biennial alternating patterns, 229E and HKU1 had peak incidence in winter seasons starting in odd number years (2009/2010, 2011/2012, etc), whereas NL63 and OC43 peaked in seasons starting in even number years (2010/2011, 2012/2013, etc).

**FIG. 2.**
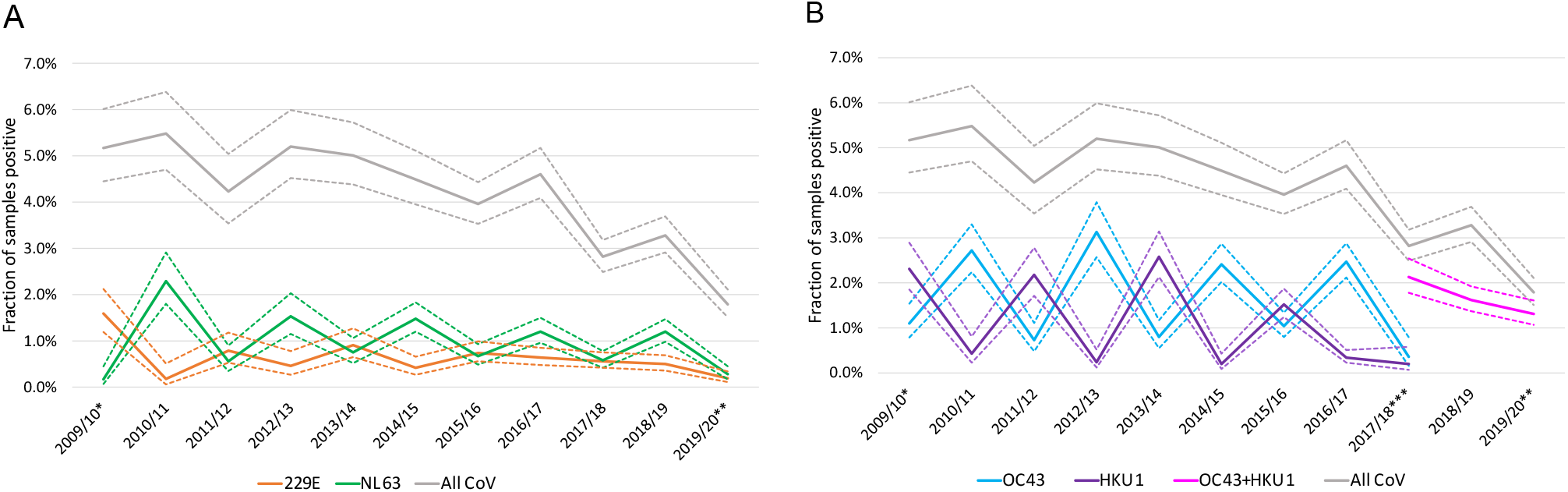
Fraction of samples per season being positive for each of the four CoV species. The fraction of samples positive per season for (A) the *Alphacoronaviruses* (229E, NL63) and (B) the *Betacoronaviruses* (OC43, HKU1). The gray line shows the fraction of samples positive for any of the CoV. Dashed lines mark CI. *, from 14 Sept until 31 Dec 2009; **, from 1 Jan until 2 April 2020; ***, from 6 November 2017 until 2 April 2020 OC43 and HKU1 were not analyzed separately.

Next, we investigated the variation of CoV detections over the calendar year by averaging over the entire study period. When all four species were combined, the lowest activity was observed from July to October. There was a steep increase in CoV detections in November, and a peak in December. However, the peaks for the individual species occurred in different months (Figure 3).

**FIG. 3.**
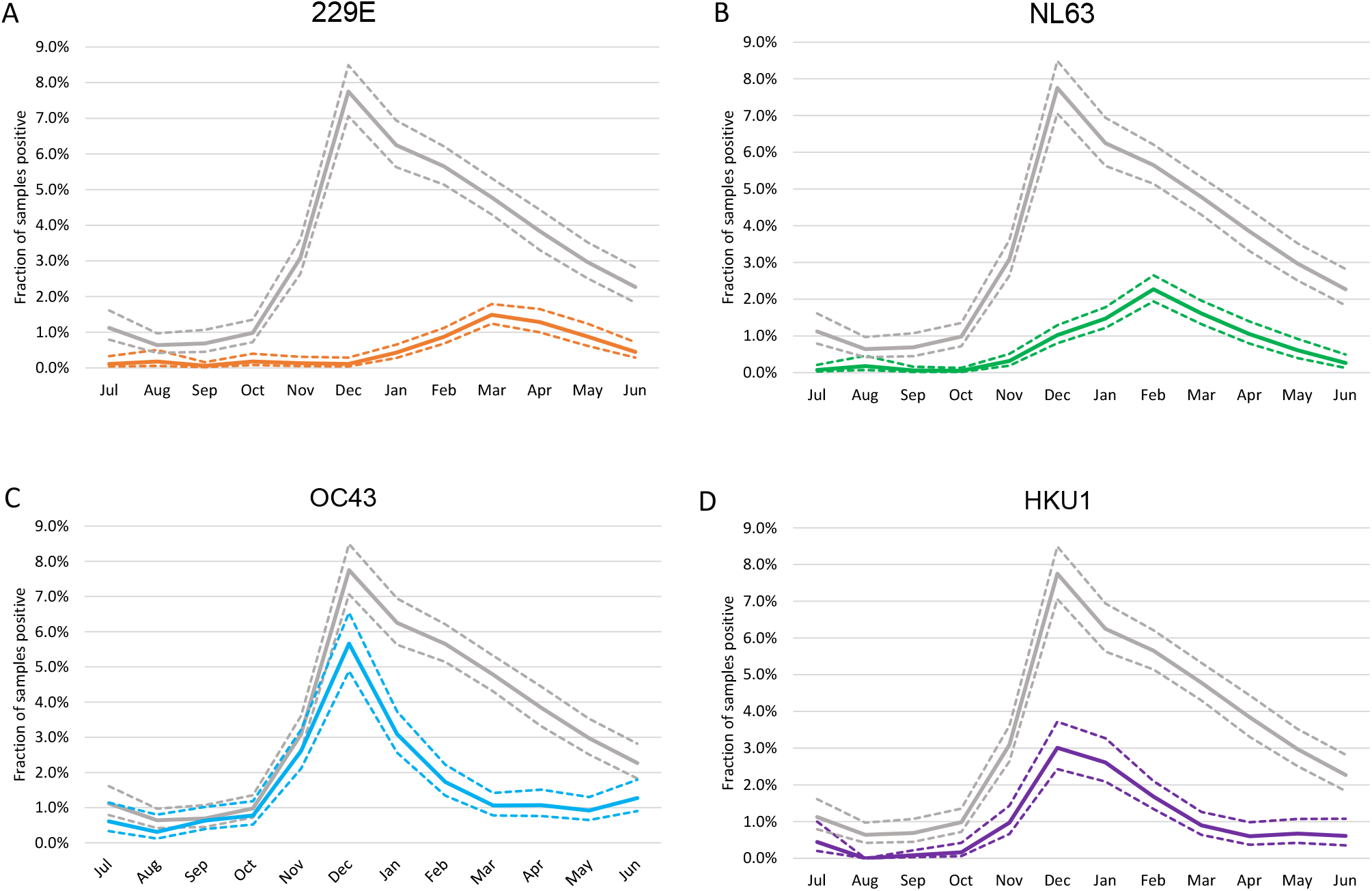
Seasonal peak. The fraction of samples per calender month being positive for (A) 229E, (B) NL63, (C) OC43, and (D) HKU1. The gray line shows the fraction of samples being positive for any of the CoV. Dashed lines mark CI.

In summary, there were two patterns in the incidence of the four species of seasonal CoVs appeared to show two different types of interactions. Firstly, both the two *Alphacoronaviruses* (229E and NL63) and the two *Betacoronaviruses* (OC43 and HKU1) peaked in alternating seasons. Secondly, the *Betacoronaviruses* peaked earlier in the winter season than the *Alphacoronaviruses*.

### Infections with seasonal CoV across age strata

Information about the age of patients with CoV-positive results was available for 55,063 (99.8%) samples. For age strata with patients younger than 20 years, more than 60% of samples had been analyzed with the extended respiratory virus panel, which included the seasonal CoVs. In contrast, less than 40% of samples were tested for CoVs in all the older age strata, and in particular the oldest patients (≥90 years) (Figure 4A). The fraction of samples positive for any CoV was highest in the two youngest age strata and was the lowest in the age stratum 80-89 years; however, it is important to note that CoV infections were diagnosed in all age strata (Figure 4A). The fractions of positive samples in the different age strata also showed some variation over seasons (supplement figures S1 and S2). We separately investigated the age distributions of each of four CoV species (Figure 4B-E, and supplement figures S1, S2). NL63 and HKU1 showed a tendency to decline with age, while detection of 229E and OC43 was more similar across the age strata.

**FIG. 4.**
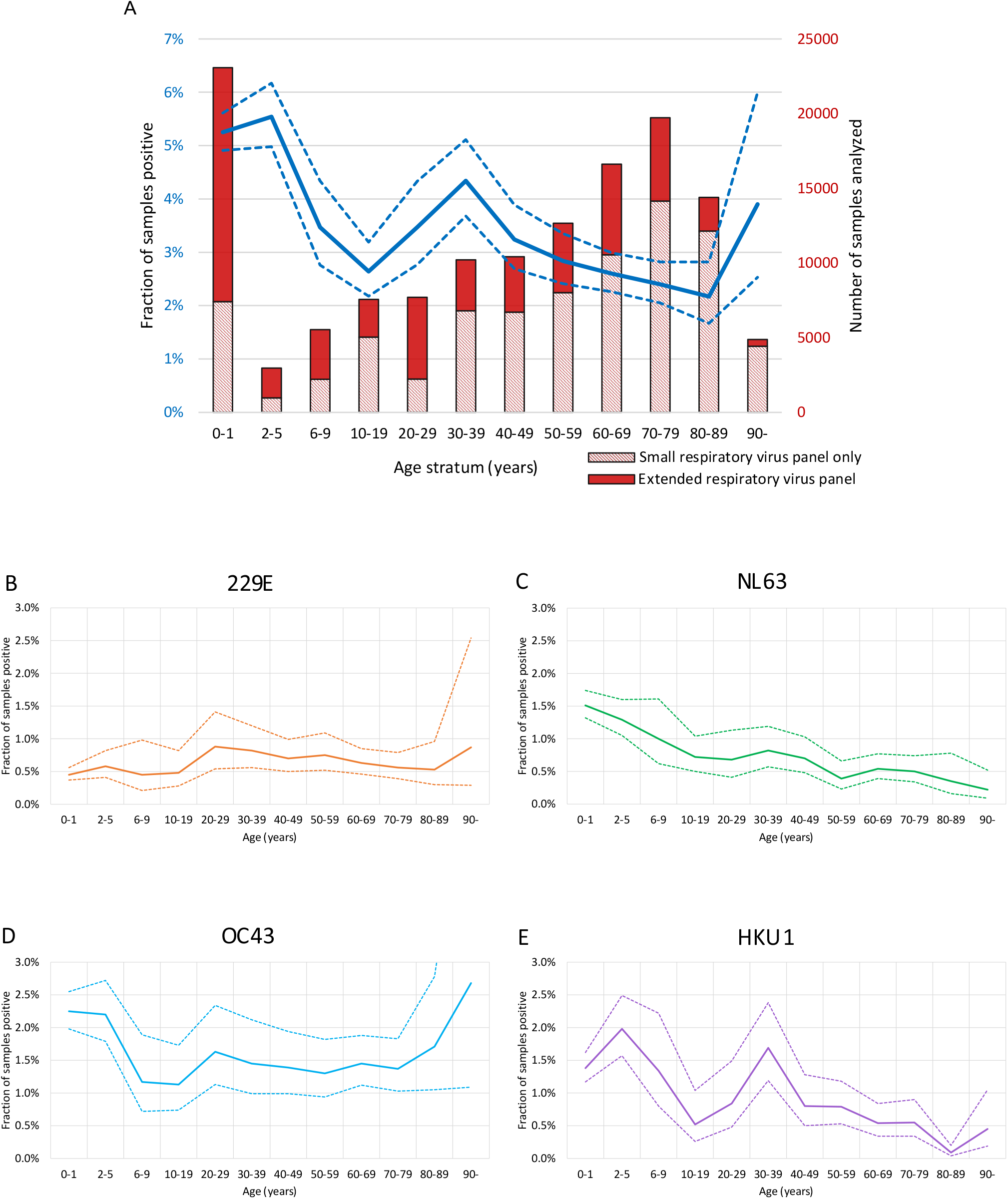
Fraction of samples positive for each species of coronavirus per age stratum. (A) Fraction of samples positive for any of the coronaviruses per age stratum (blue line, dash lines mark 95% CI). Bars show number of submitted samples in total (*n* = 135,922), and fraction analyzed for the extended respiratory panel which includes the analysis for coronaviruses (solid red, *n* = 55,190). Fraction of samples positive per age stratum for (B) 229E, (C) NL63, (D) OC43, and (E) HKU1.

### Co-detection of CoVs with other respiratory viruses

Of 2,130 samples positive for any CoV, matched results for analyses of the full panel of other respiratory viruses were obtained for 1,953 samples. At least one additional respiratory virus was identified in 737 samples (34.6%, 95% CI: 32.6-36.7%), and two or more other respiratory viruses were identified in 128 samples (6.0%, 95% CI: 5.0-7.1%). Co-detections were most common in the two youngest age strata (0-1 and 2-5 years), accounting for 76.1% of samples positive for CoV and a co-detected virus. The four viruses that were most commonly co-detected with CoVs were: RSV (9.2%), RV (7.2%), BoV (6.8%) and AdV (4.7%) (Figure S3); the co-detections appeared less common than expected by chance. Thus, the odds ratio was significantly reduced for co-detection of CoVs with RSV (0.56, 95% CI 0.47 to 0.66) and AdV (0.52, 95% CI 0.42 to 0.66), and non-significantly reduced for RV 0.83 (95% CI 0.62 to 1.12) and BoV 0.95 (95% CI 0.79 to 1.15). However, more than half (59.4%) of the samples were submitted for analysis IAV, IBV, and RSV only. In these samples, IAV was detected in 14.6% of samples, compared to detection of IAV in 4.6% of samples with request for the extended respiratory virus panel. Thus, co-detection of the seasonal CoVs with RSV and the influenza viruses might be underrepresented in this material due to selection bias.

## Discussion

In this study we comprehensively have investigated the epidemiology of the four seasonal coronaviruses (229E, NL63, OC43, and HKU1) using PCR results from more than 55,000 clinical samples analyzed 2009–2020 in Stockholm, Sweden. We found that CoV infections were detected in around 4% of samples, but due to considerable seasonal variation the proportion positive samples varied from <1% in Aug–Sept, to close to 8% in Dec–Jan. All four seasonal CoVs showed biennial winter incidence peaks, with alternating peak seasons for the two *Alphacoronaviruses*, 229E and NL63, and the two *Betacoronaviruses*, OC43 and HKU1, respectively. This novel finding suggests some type of immunological interaction or interference. The fraction of CoV-positive samples was the highest among children aged 0-5 years, but infections occurred in all age strata, which suggests that symptomatic reinfections among adults and the elderly are not uncommon. Our results concerning the epidemiology of seasonal CoVs have implications for the likely future endemic presence of SARS-CoV-2 and extend the knowledge gained by earlier studies [9-11].

We report that the four species of seasonal CoVs appeared to show two different types of interactions. Firstly, both the *Alphacoronaviruses* (229E and NL63) and the *Betacoronaviruses* (OC43 and HKU1), respectively, peaked in alternating winter seasons. Secondly, the circulation of the *Betacoronaviruses* peaked earlier in the winter season than that of the *Alphacoronaviruses*. The biennial pattern of the seasonal CoVs that we report is in line with earlier reports from the temperate zone of the Northern Hemispere [1, 4, 11-13]. Importantly, these studies also found that *Alphacoronaviruses* and *Betacoronaviruses*, respectively, tended to peak in alternating seasons, matching our findings. However, only Kissler *et al*. [11] and Nickbakhsh *et al*. [9] make a point of this finding and draw inference about possible immunological interactions and their implications for the future of SARS-CoV-2, but the studies did not include *Alphacoronaviruses* or HKU1, respectively. Thus, we extend their findings by showing that the both genera show similar interactions, which indicates that this is a general property of CoVs. Furthermore, it is reassuring that the biennial alternating pattern is found in different continents and countries in the temperate zone of the Northern Hemisphere, as this indicates that the interactions are not a local phenomenon in our study setting in Stockholm, Sweden. It should be noted that the effect may be limited to the temperate zone as biennial seasonality is not noted in a study from southern China [2].

The alternating biennial cycles of the two *Alphacoronaviruses* and the two *Betacoronaviruses* warrants further investigation. It could be a chance event, but more likely is due to some type of anti-phase synchronization [31], which could be generated even by weak cross-immunity. With regard to immunity, although IgG antibodies against CoVs are present in almost all adult individuals [14], early studies showed that CoV infections commonly occur despite a prior presence of neutralizing antibodies [13, 15, 16]. Moreover, a substantial number of reinfections of NL63 were detected in a recent study with repeated sampling over a season in community setting [17]. Indications of frequent reinfections with all four seasonal coronaviruses were also provided by a recent serological study from the Netherlands [18], as did our study, with infections occurring in all age groups. Immunologic interaction between the seasonal CoVs has been suggested as a cause of the dominance of NL63 and OC43 in infants [19].

The prevalence of seasonal CoV infection worldwide and possibility of immunological interference by seasonal CoVs with each other has raised hopes of potential protective effects against SARS-CoV-2-infection or modulation of COVID-19 disease [20]. However, despite the presence of cross-reactive binding antibodies between SARS-CoV and SARS-CoV-2, cross-neutralization appears to be rare [21], even though these viruses are relatively closely related. All of these findings are interesting in the context of the SARS-CoV-2 pandemic, because T-cells reactive to SARS-CoV-2 have been detected in samples from donors sampled before the pandemic [22, 23], as well as in seronegative exposed persons [24]. This could possibly be due to cross-immunity between the seasonal CoVs and SARS-CoV-2 [9, 11].

As we find a higher prevalence of seasonal CoV infections in younger age groups, recent seasonal CoV infection and some level of cross-reactive immunity with SARS-CoV-2 might at least partially explain the apparently lower attack rate of SARS-CoV-2 in young children compared to older persons [25, 26]. At the same time, reinfection by seasonal CoVs clearly occurs throughout life and any potential cross-protection might be short-lived [18], implying that the magnitude of the cross-protective effect might be small.

We found that OC43 was the species that was most commonly detected, which is in line with earlier studies [1-4, 9, 12, 27, 28]. The odds ratio for positive samples in our study was significantly lower for females than males. This gender difference is interesting in relation to COVID-19, as male patients have a higher risk of severe disease and death than females [29, 30]. We noted CoV infections across all age strata, although the highest prevalence was observed among children. Collectively, our results indicate that symptomatic CoV reinfections among adults and elderly are not uncommon, though we did not formally exclude the possibility that they had primary CoV infections through serological testing.

Our study has some limitations. In particular, it is a retrospective study that utilizes results from routine clinical diagnostics. It is difficult to exclude that there have been changes in the strategies and prioritization for diagnostics of respiratory infections over the study period. For example, there have been some changes in the platforms used for virus diagnostics during the study period that may have modified the sensitivity of CoVs detection. Also, information about clinical presentation and disease severity were not available. However, all samples were from clinical diagnostics, which means that the symptoms were severe enough to prompt sampling. Another limitation is that more than half of the adults were tested only for IAV, IBV, and RSV, and not the full virus panel. It is likely that CoV infections have been underdiagnosed among the adults and the elderly because they were not tested for. The main strengths of the study are the large sample size and the long study period.

To conclude, the notable yearly alternation within genera, and the temporal spacing in peaks between genera, suggest a possible immunological interaction or interference. We further identify a distinct difference in the timing of the seasonal peaks, with the *Alphacoronaviruses* peaking months after the Dec-Jan peak observed for the *Betacoronaviruses*. Our findings have implications both for better understanding seasonality and interaction in coronaviruses generally and for the likely future endemic presence of SARS-CoV-2, and thus merit further immunological studies.

## Data Availability

Data available within the article or its supplementary materials.

## Acknowledgements

We thank Berit Hammas, MD, at the Department of Clinical Microbiology, Karolinska University Hospital, for valuable input on the virus diagnostics.

## Transparency declaration

The authors declare no conflicts of interest.

## Authors’ contribution

Concept and design: RD, EBH, RAN, JA.

Analysis and interpretation of data: RD, EBH, MW, RAN, JA. RD, MW, and JA accessed the underlying data.

Drafting of the manuscript: RD, JA.

Critical revision of the manuscript for important intellectual content: RD, EBH, MW, RAN, JA.

All authors finally approved the version to be submitted.

