## Supplement for "Interactions between seasonal human coronaviruses and implications for the SARS-CoV-2 pandemic: A retrospective study in Stockholm, Sweden, 2009-2020"

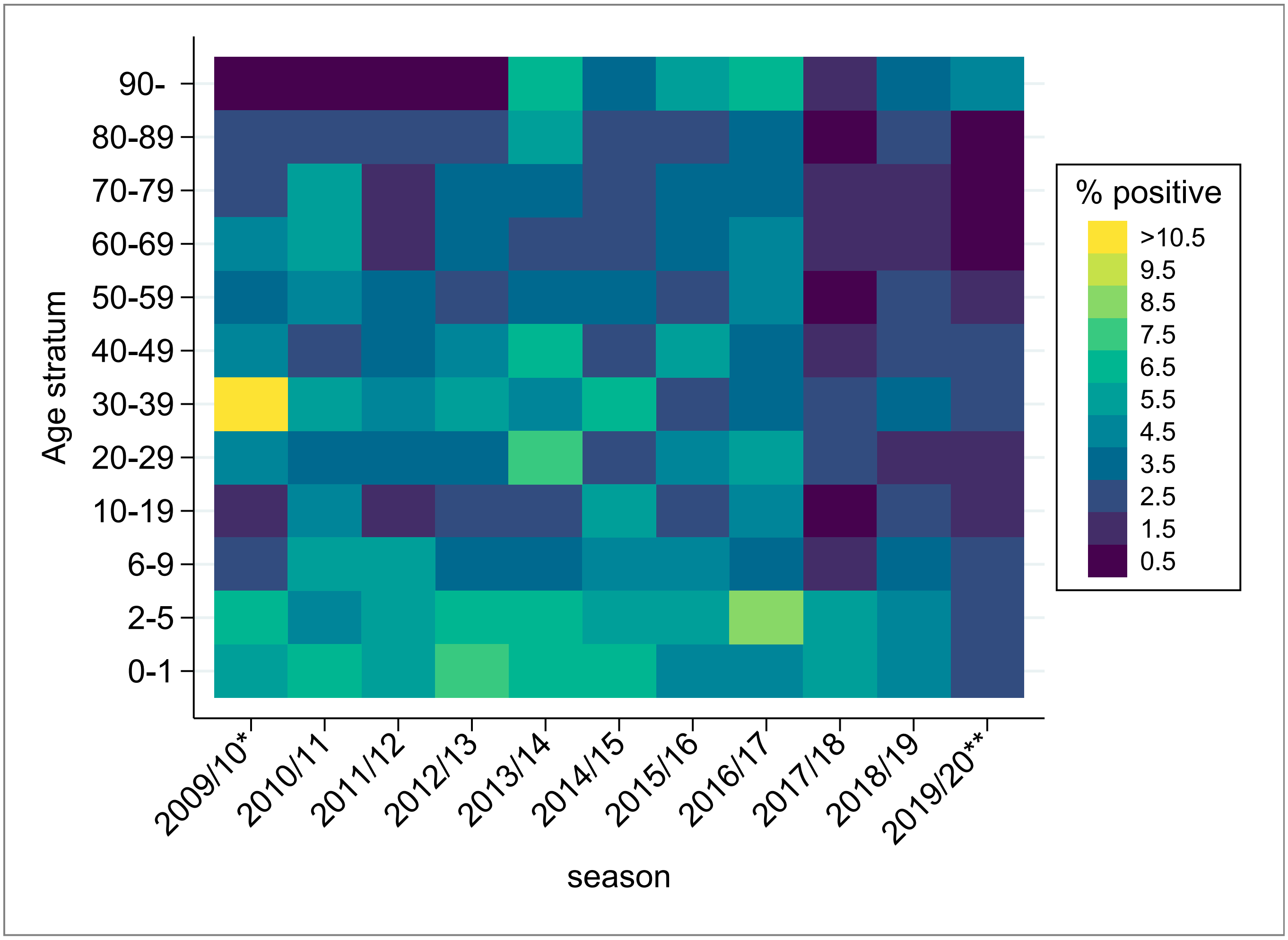


FIG. S1. **Positive fraction per age stratum**. The heatmap shows the fraction of samples per age stratum being positive for any of the four seasonal coronaviruses.
*, from 14 Sept until 31 Dec 2009; **, from 1 Jan until 2 April 2020.

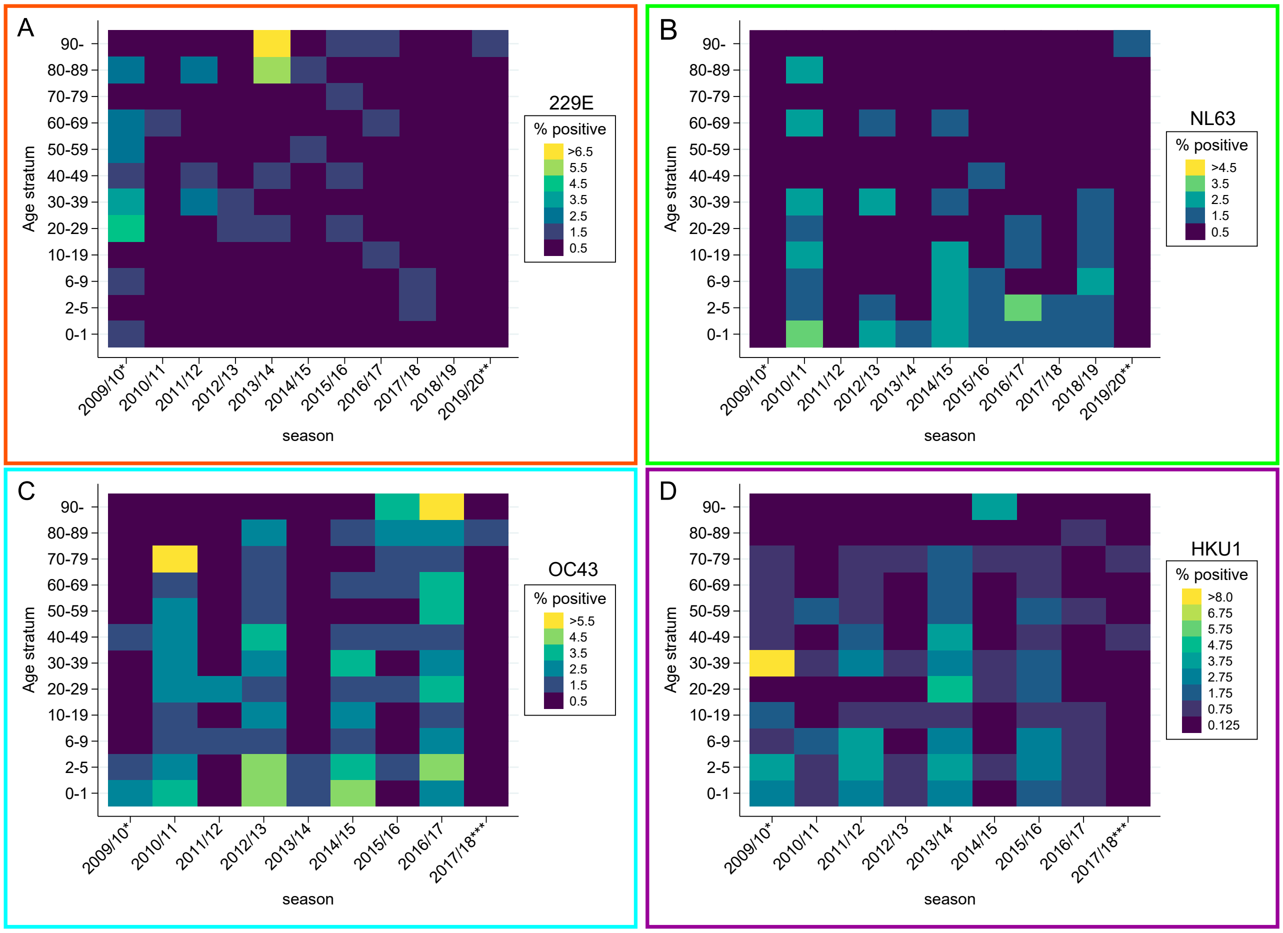


FIG. S2. **Positive fraction per age stratum, per species**. The heatmaps shows the fraction of samples per age stratum being positive for (A) 229E, (B) NL63, (C) OC43, and (D) HKU1.
*, from 14 Sept until 31 Dec 2009; **, from 1 Jan until 2 April 2020; ***, from 1 Jan until 5 Nov 2017.

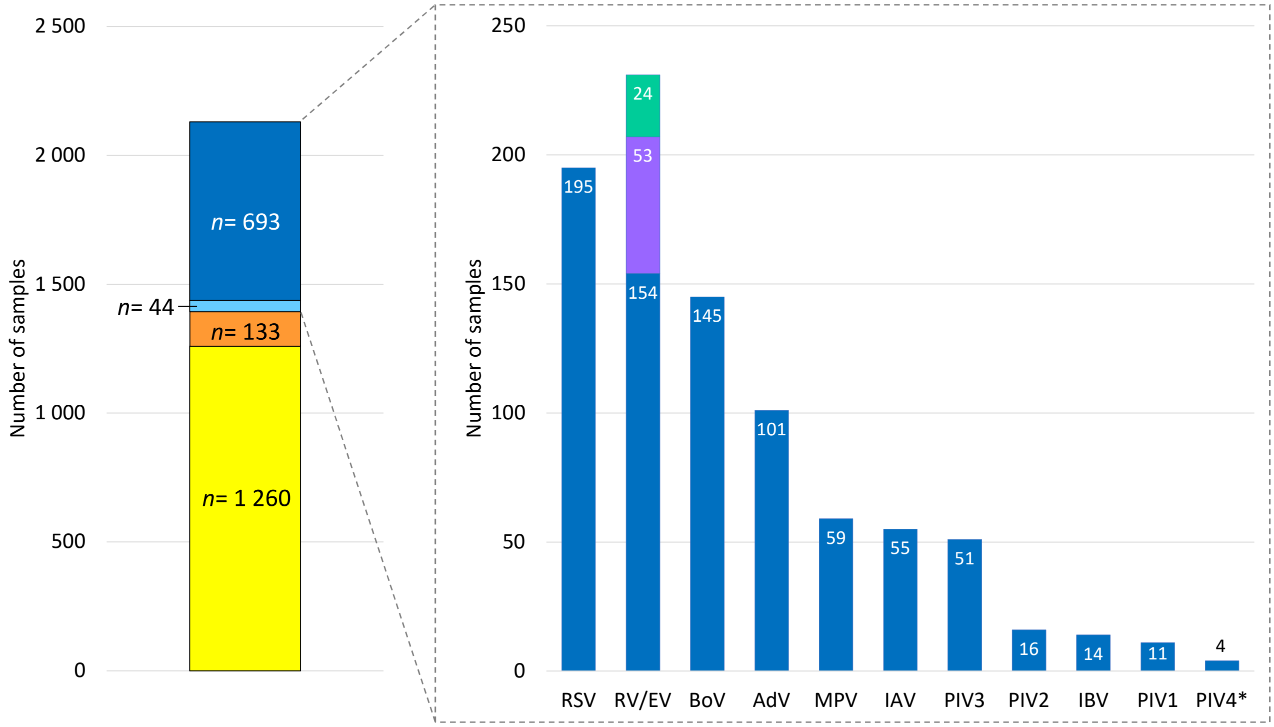


FIG. S3. **Co-detections.** Left bar: stacked in bar bottom to top ($n$ total = 2,130): mono-detection of CoV ($n$=1,393) (complete records: yellow; missing results for narrow respiratory panel: orange); and CoV with a co-detected respiratory virus ($n$=737) (missing records for narrow respiratory panel (IAV, IBV, and RSV): light blue; complete records: dark blue). Insert at right: Co-detected pathogens in samples positive for CoV. Bar RV/EV, bottom to top: mono-detection of RV (dark blue); dual detection RV and EV (purple); mono-detection EV (green).
*, PIV4 analyzed only from 6 Nov 2017 until 2 April 2020.
